# Escaping the Negative Attentional Bias in Depression with Real-Time Neurofeedback

**DOI:** 10.64898/2026.09.30.26363896

**Authors:** Nitzan Lubianiker, Brendan Woods, Frederich Nitchie, Alexandra Batzdorf, Anne Mennen, Qi Lin, Kenneth A. Norman, Nicholas B. Turk-Browne, Yvette I. Sheline

**Affiliations:** Department of Psychology, Yale University, New Haven, CT, USA; Princeton Neuroscience Institute, Princeton University, Princeton, NJ, USA; Center for Neuromodulation in Depression and Stress, Department of Psychiatry, Perelman School of Medicine, University of Pennsylvania, Philadelphia, PA, USA; Center for Neuroscience Imaging Research, Institute for Basic Science, Suwon, South Korea; Center for Brain Science, RIKEN, Wako, Japan; Department of Psychology, Princeton University, Princeton, NJ, USA; Wu Tsai Institute, Yale University, New Haven, CT, USA

**Keywords:** Brain-Computer Interface, Major Depressive Disorder, Selective Attention, fMRI

## Abstract

Individuals with major depressive disorder (MDD) show an attentional bias toward negatively valenced stimuli and thoughts. In this study, we applied a closed-loop neurofeedback procedure designed to reduce this bias. Participants were shown composite negative faces and neutral scenes with variable opacity and were instructed to attend to the neutral scene while ignoring the negative face. Internal attentional states were decoded in real time from functional magnetic resonance imaging (fMRI) data. When a participant’s decoded attentional state indicated a failure to ignore the negative faces, the faces became more visible (higher opacity), thus externalizing the brain’s attentional lapse in that moment. Forty-eight individuals with MDD were randomly assigned to a real neurofeedback training group (N = 24) or a sham control group (N = 24); the control group received feedback yoked to a participant in the real group. All participants completed three fMRI neurofeedback sessions. The main outcome quantified the extent to which participants got “stuck” in the most negative attentional state of focusing strongly on the negative faces. We hypothesized that the real neurofeedback group would learn to escape that state by the end of training, relative to the sham control group. Consistent with our hypothesis, neurofeedback training reduced the probability of getting stuck in the most negative attentional state. In offline analyses, training reduced fMRI activity in the precuneus/posterior cingulate and the medial prefrontal cortex when participants successfully attended to scenes and ignored faces. These results demonstrate the efficacy of remediating the negative attentional bias in depression with closed-loop neurofeedback from real-time fMRI.

## Introduction

We face considerable emotional challenges in everyday life, ranging from external threats and stressful social interactions to internal negative thoughts. Attention helps address these challenges: we can attend to valenced stimuli to process and respond to them, and then disengage when attending is no longer beneficial. Individuals with major depressive disorder (MDD) struggle to deploy their attention flexibly in this manner; instead, they show an inflexible attentional bias to-wards negative stimuli and thoughts (1). Once they attend to negative content, they struggle to disengage from those states (2–4), leading to a self-reinforcing feedback loop that worsens mood, clinically manifested as cognitive rumination (5).

Standard behavioral therapy approaches (e.g., attention bias modification training) have relatively weak efficacy in treating this bias (6, 7), arguably because they try to modify the behavioral outputs of maladaptive neural processing patterns rather than the underlying neural mechanisms themselves. A recent proof-of-concept study employed a functional Magnetic Resonance Imaging (fMRI) neurofeedback training aimed at remediating this bias by targeting its underlying neural mechanisms (8). This study compared the effects of neurofeedback training on MDD patients and healthy controls, and found that neurofeedback significantly improved the ability of MDD patients to disengage from negative stimuli relative to healthy controls.

Two critical questions remain unaddressed by prior work. First, are improvements in disengagement from negative states specific to participants receiving neurofeedback or do they reflect non-specific placebo effects, which are known to be substantial in MDD (9, 10)? To answer this question, it is necessary to compare a group of MDD individuals receiving neurofeedback based on their own attentional state to an MDD control group that receives sham neurofeedback that is not contingent on their own attentional state. The proof-of-concept study mentioned above did not include an MDD control group and thus cannot rule out placebo effects. Second, what are the neural consequences of neurofeedback training? MDD is characterized by distinct patterns of emotional information processing, and it is unclear whether neurofeedback training improves attentional disengagement by altering these maladaptive patterns.

In the current study we used a double-blind, randomized controlled design with a yoked sham control group to test how fMRI neurofeedback training reduces negative attentional biases beyond the non-specific improvements typical of depression treatments (9). The yoked sham control matches instructions, task demands, stimuli, and reward, but crucially these participants receive feedback based on a previous neurofeedback participant’s brain activity. This allows us to test whether feedback contingent on one’s own brain activity is required for beneficial effects in this paradigm. We then examined how such training influences neural mechanisms of attention to negative stimuli in MDD.

## Results

Each training run contained multiple blocks of a go/no-go task in which translucent face and scene stimuli were superimposed into a composite image, with instructions that varied across blocks (**Figure 1A**). During the first run and the first four blocks of each of runs 2-9, participants were instructed either to attend to the location of every scene (e.g., indoor: go; outdoor: no-go) while ignoring neutrally valanced faces, or to attend to the sex of every face (e.g., male: go; female: no-go) while ignoring the scenes. Throughout these blocks, face and scene opacity remained constant (50% each), so that both image categories were equally visible. Brain responses during blocks when participants attended to the scene or attended to the face were used to train multivoxel pattern classifiers to distinguish scene and face processing in the brain (**Figure 1B**). During the subsequent neurofeedback blocks (blocks 5-8) of runs 2-9, participants were instructed to attend to the scene and perform the indoor/outdoor go/no-task, while ignoring the faces that were now negatively valenced. In these blocks, trained classifiers controlled the face and scene opacity: when a participant’s classifier indicated a failure to ignore the negative faces, the faces became more opaque (i.e., more visible) and the scenes more transparent. This externalized the brain’s off-task attentional state at the moment the bias toward negative content was expressed, motivating participants to refocus attention on the neutral scenes. Conversely, when the classifier indicated scene processing (consistent with the instructions), the faces became more transparent and the scenes more opaque. This regime essentially rewarded participants for not attending to the negative face by making the scene judgment task easier, thus associating reward with successful disengagement from negative content.

**Fig. 1.**
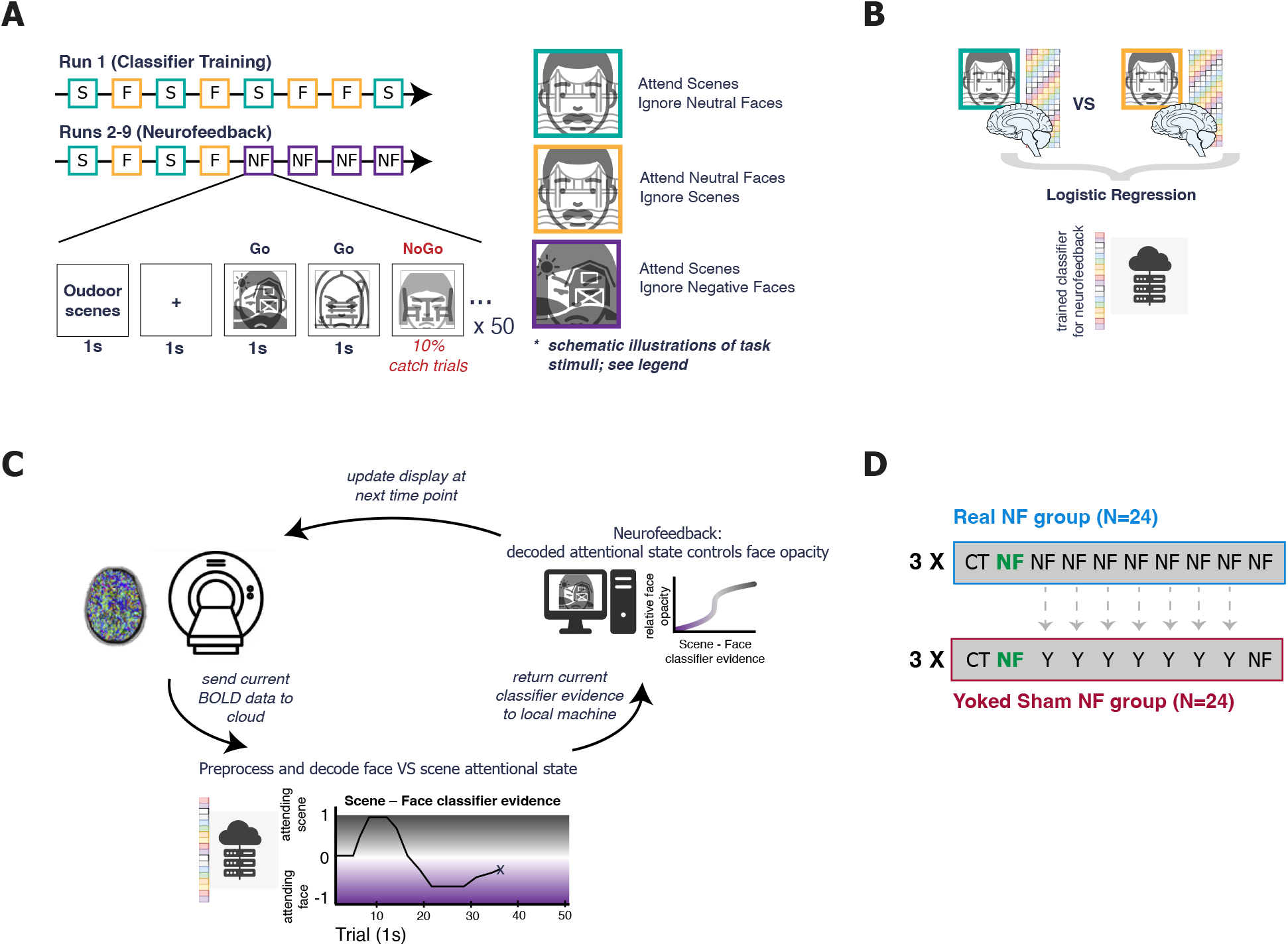
Study Design. A. Training included runs containing blocks of a Go-NoGo task with 50 superimposed face and scene images. During all of run 1 (classifier training) and the first four blocks of runs 2-9, instructions alternated between attending to the location of the scene (indoor/outdoor) and ignoring neutral faces (turquoise), and attending to the sex of the face (male/female) and ignoring scenes (yellow). During the last four blocks of runs 2-9 (neurofeedback, NF; purple), the faces were now negative and participants were instructed to ignore them and attend to the scene. Face and scene stimuli are depicted schematically for illustration only; examples of the original stimuli can be found in (8). B. Before neurofeedback began (between blocks 4 and 5 in runs 2-9), blood oxygenated level-dependent (BOLD) data from the previous 8 blocks displaying neutral faces were sent to a cloud computer, where logistic regression pattern classifiers were trained to decode attention to scene vs. face categories. These classifiers were tested during the subsequent neurofeedback blocks to decode face vs. scene attention. C. Overview of neurofeedback set-up using RT-Cloud software (11). At each time point (TR = 2 s), an acquired BOLD volume was masked and sent to a remote cloud server, where it was pre-processed, after which the trained pattern classifier decoded the extent to which attention was directed at the task-relevant scene or the task-irrelevant face. To yield a single signed feedback signal (positive toward the scene, grey; negative toward the face, purple), the difference between classifier evidence for the scene (ranging from 0 to 1) and face (ranging from 0 to 1) was calculated; an example time course is illustrated in the lower panel. This difference was sent back to the task computer at the MRI to influence the appearance of the display the participant viewed during the following time point. Classifier output was converted to opacity level via a sigmoidal function. When attention was drawn toward the task-irrelevant negative faces, the faces became more opaque, motivating participants to refocus attention back to the task-relevant scenes. D. Each participant completed three training sessions with nine runs each. During runs 3-8, each control participant from the yoked sham group saw the time-varying face opacity levels of a matched participant from the real neurofeedback group instead of their own decoded attentional states (yoked feedback is indicated with a Y in the diagram). The first (run 2) and last (run 9) runs provided genuine neurofeedback for yoked participants as well so we could obtain our main outcome measure of the participant’s ability to escape negative attentional states. The main analyses focused on run 2 (green) in both groups, which provided a clean assessment of attentional abilities near the beginning of each session while participants were fresh.

### Negative attentional biases

The main outcome, measured in the first and last neurofeedback runs in each session, quantified the extent to which participants got “stuck” in the most negative attentional state. This occurred because of a feedback loop: when participants attended to the negative face (despite being task-irrelevant), it became more opaque as neurofeedback, this greater opacity increased attention to the face, making it more opaque, and so on. With only a faint scene remaining in the image, participants struggled to draw their attention back to the task-relevant neutral scene. We predicted that neurofeedback training would help them learn how to get unstuck, a skill that could carry over to mitigate the negative attentional bias by helping individuals with MDD self-modulate their attention toward neutral stimuli.

To compare attentional states across time, we divided the continuous scene vs. face classifier evidence into 16 bins (**Figure 2A**, top panel). Then, we compared the attentional state at time t to the attentional state at time t + 5 seconds (when we would expect feedback effects on brain activity to be maximal, accounting for the hemodynamic lag). As each TR was 2 seconds, we calculated the attentional state at t plus 5 seconds by averaging the data from 2 and 3 TRs after t. Specifically, for each time delay d (2 or 3 TRs), for a given attentional state bin A, we calculated the persistence of that state by counting the number of times that the scene minus face classifier evidence fell within A at time t + d; we then divided this number by the total number of occurrences of state A to normalize to a conditional probability we call “stickiness” henceforth (**Figure 2A**, bottom panel). Our hypothesis was that the real neurofeedback group would get better at escaping the most negative attentional state from early to late in training, and thus show reduced stickiness for the most negative attentional state bin, relative to the sham control group.

**Fig. 2.**
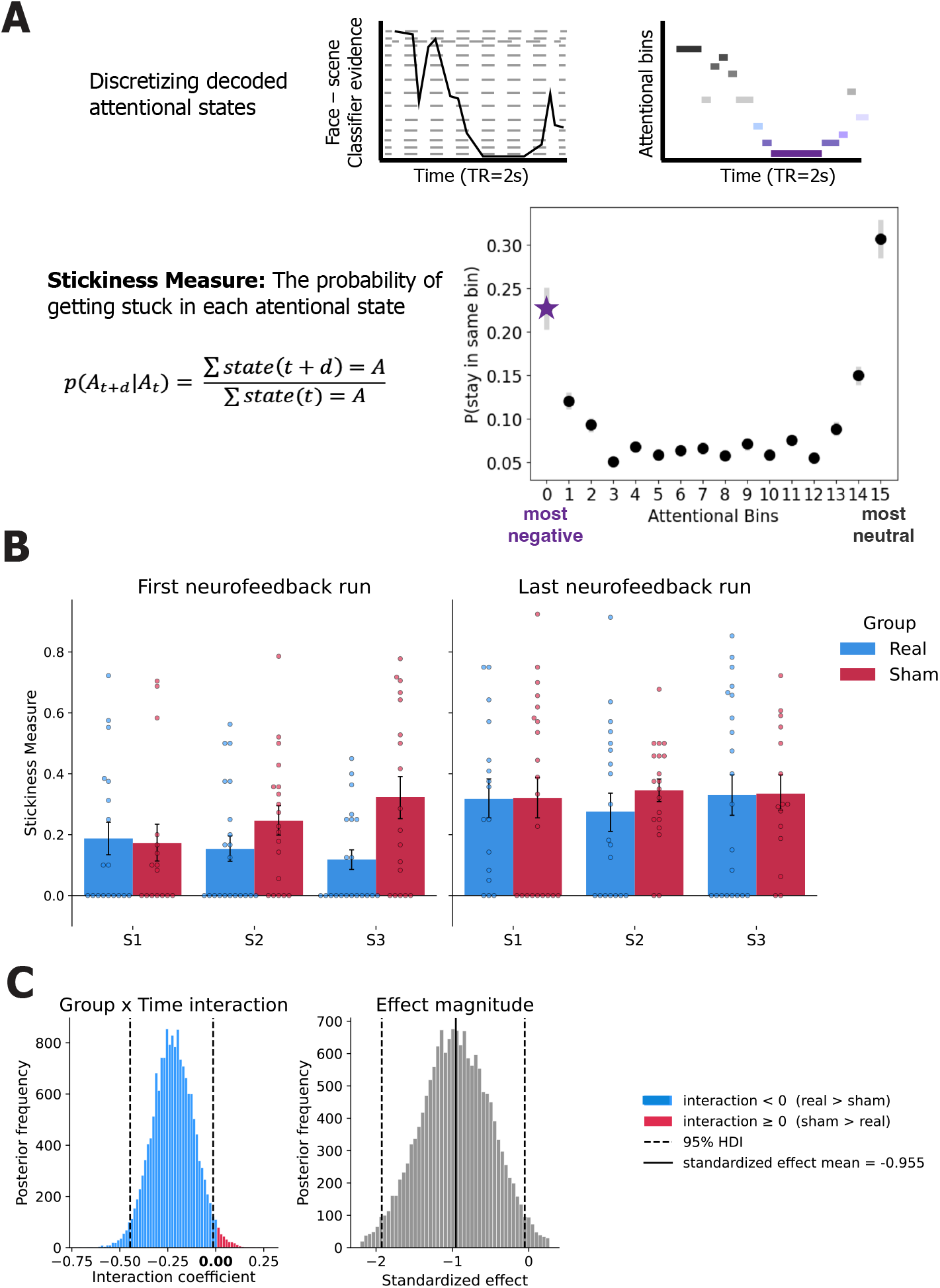
Training effects. A. Stickiness measure calculation. First, the continuous face-vs.-scene classifier evidence per run were divided into 16 attentional bins, so that each time point is characterized by a discrete value ranging from the most neutral state (fully attending to scene, black) to the most negative state (fully attending to the negative face, purple). The upper panel shows an example transformed time course. Next, for each attentional bin, we calculated the probability of getting stuck in that state. The lower panel shows the grand mean of stickiness per attentional bin collapsed across groups and sessions. The preregistered outcome measure was the stickiness for the most negative attentional bin (marked by a purple star). B. Means and standard errors of the mean of the stickiness measure across groups (blue: real NF, crimson: sham NF) and sessions (1 to 3) for the first (run 2, left panel) and the last (run 9, right panel) neurofeedback runs. Dots show individual participants’ stickiness scores. C. Differences between groups in stickiness across sessions for run 2, fit with a Bayesian linear mixed effects model. Left panel: the posterior distribution of the interaction coefficient, capturing the extent to which stickiness in the real group decreased from session 1 to 3 compared to the sham group. Blue bars indicate larger decreases in the real group (consistent with our hypothesis), and crimson bars correspond to larger decreases in the sham group. Dashed lines indicate the 95% HDI. Right panel: Posterior distribution of the standardized effect of group-by-session interaction (interaction coefficient/sigma) estimating effect magnitude. The horizontal black line marks the distribution mean, and the dashed lines mark the 95% HDI.

**Figure 2B** presents the means and standard errors of the mean per across runs, sessions, and groups. There was an overall increase in stickiness from the first to the last run of each session (run 2 mean = 0.20, run 9 mean = 0.32; paired *t*(46) = *−*3.50, *p* = 0.001, Cohen’s *d* = *−*0.51). We interpret this finding as a fatigue effect that grew toward the end of sessions, consistent with prior fMRI neurofeedback studies (12) and sustained attention tasks (13). Because these generic, within-session fatigue effects could potentially mask effects of training, we focused stickiness analyses on the first neurofeedback run in each session (run 2), when participants were fresh, to assess training-related differences across sessions and groups.

Two characteristics of the data influenced our choice of statistical model for estimating group-by-session training effects. First, the data were not distributed normally, with the stickiness measure bounded [0, 1] and skewed towards zero. Stickiness distributions for both sessions 1 and 3 were zero-inflated (47% and 44% of values equal to zero, respectively) and left-skewed (session 1: skewness = 1.15, D’Agostino skewness test *z* = 2.75, *p* = 0.006; session 3: skewness = 0.89, *z* = 2.35, *p* = 0.019). Second, there were a number of missing data points. This is because of how the stickiness measure is computed: if a participant does not visit the most negative attentional bin during a run, the stickiness measure for that bin remains undefined. Indeed, only 16 real and 12 sham participants were with measurements in run 2 for both sessions 1 and 3.

We thus employed a Bayesian hierarchical mixed linear model (14). With a random intercept per participant, it estimated the group-by-session interaction without relying on normality, basing inference on the full posterior (a directional probability and 95% Highest Density Interval, HDI) rather than a single test statistic. Critically, it models missing stickiness values under the likelihood rather than requiring complete cases, retaining 23 real and 22 sham participants with at least one of the two timepoints. Nonetheless, the results below replicated in the 16 real and 12 sham participants who had complete data via a Wilcoxon rank-sum test and using a non-parametric bootstrap resampling analysis (see Supplementary Information).

The Bayesian model included fixed effects for group (real, sham), session (1, 3), and their interaction, a random intercept per participant, weakly informative priors (Normal(0, 1) on the fixed effects; Half Normal(1) on the participant and residual standard deviations), and partially observed values modelled under the likelihood so that 45 participants with at least one timepoint were retained (23 real, 22 sham). The posteriors were sampled with NUTS (4 chains, 2,000 tuning and 4,000 draws each; 16,000 posterior samples). The interaction coefficient mean was negative, and the 95% (two-tailed) HDI upper bound was lower than zero, indicating a greater pre-to-post reduction in stickiness in the real compared to the sham group (posterior mean of interaction *β* = −0.223; SD = 0.110, 95% HDI [−0.446, −0.014]; **Figure 2C**, left). The standardized effect distribution (*β* interaction/*σ*) estimating effect magnitude (mean = −0.955, 95% HDI [−1.925, −0.052]), corresponds to a moderate-to-large effect (**Figure 2C**, right). Sampling diagnostics were satisfactory for the effect of interest (*R* = 1.00, bulk ESS *≈* 7, 900; 1 divergence out of 16,000). These results support our hypothesis that real neurofeedback training, relative to a sham control, can reduce the negative attentional biases in MDD.

### Neural changes

In offline fMRI analyses, we explored whether reducing the attentional bias also altered the evaluative and attentional circuitry engaged by the training task. We modeled how the fMRI signal differed during periods where scenes were more visible (reflecting on-task performance) vs. less visible (reflecting off-task performance) and then computed how this response changed from early to late in training. Specifically, we modeled the parametric effect of scene vs. face visibility on the BOLD signal during the neurofeedback blocks and contrasted the change from sessions 1 to 3 between groups. Two participants were excluded from these analyses because of excessive head movements (see Methods), leaving 24 real and 22 sham participants.

A whole-brain analysis using a randomization test with threshold-free cluster enhancement (TFCE; *p <* 0.05 familywise error, FWE) revealed significant group differences in two distinct clusters: at the intersection between the precuneus and posterior cingulate cortex (PCC) (**Figure 3**, top row) and the medial prefrontal cortex (mPFC) (**Figure 3**, bottom row). This result indicates that from early to late in training, real compared to sham neurofeedback was associated with decreased activity in both clusters when participants were successfully attending to scenes and ignoring faces.

**Fig. 3.**
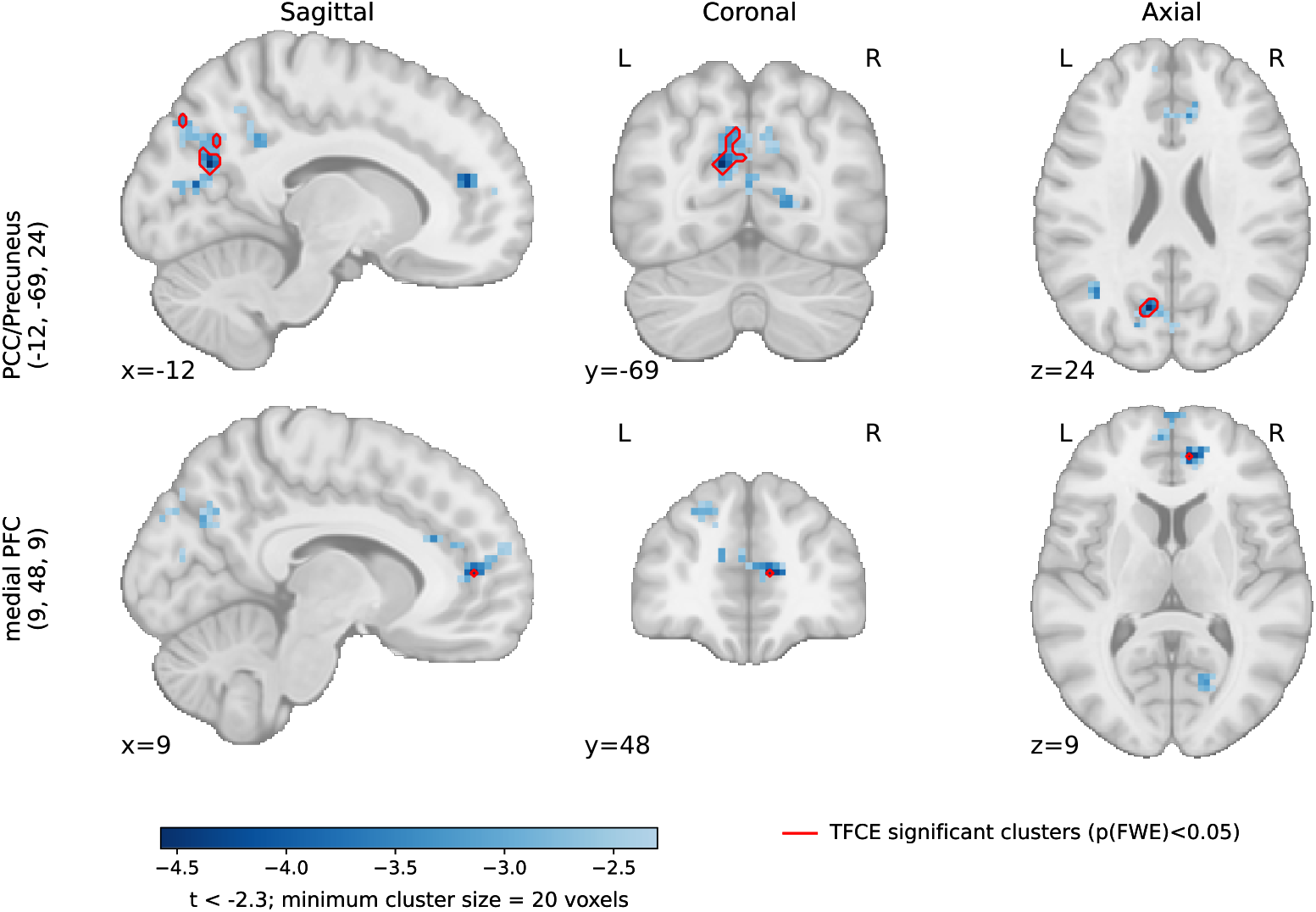
Whole-brain training-related changes in the parametric fMRI response to scene vs. negative face visibility (real vs. sham, session 3 vs. session 1). Top row: posterior cingulate cortex (PCC)/precuneus (peak coordinate in standard Montreal Neurological Institute space: −12, −69, 24). Bottom row: medial prefrontal cortex (PFC) (9, 48, 9). Sagittal, coronal, and axial views are shown for each cluster at the indicated slice coordinates. The *t* statistic values (displayed at *t < −* 2.3, minimum cluster size 20 voxels) are denoted by a blue color scale; red outlines indicate clusters surviving TFCE correction at *p*(FWE) *<* 0.05. Negative values indicate training-induced activity decreases while participants were successfully attending to scenes and ignoring faces.

Next, because the amygdala’s role in processing negative faces is shaped by its connectivity with regulatory regions and face-selective visual cortex (15), we explored whether training reshaped task-modulated connectivity between these regions. We used a parametric psychophysiological interaction (PPI) with a left-amygdala seed and scene vs. face visibility as the psychological factor. We did not find reliable task modulation of connectivity between the left amygdala and a fusiform face area (FFA) ROI (*p* = 0.682). We also performed this analysis across the whole brain but no clusters showed task-modulated amygdala connectivity that survived correction for multiple comparisons.

### Transfer effects

A secondary goal of this study was to test whether training effects transferred to other behavioral and clinical contexts. To assess behavioral transfer, participants performed a free-viewing gaze task measuring gaze allocation toward stimuli of different emotional valence (fearful, sad, positive, or neutral), administered at the first visit and after each neurofeedback session. To assess clinical transfer, participants completed the Montgomery–Åsberg Depression Rating Scale (MADRS; Montgomery & Åsberg, 1979) after the final neurofeedback session and again at 1-week and 1-month follow-ups. We found no significant group differences on either the behavioral or the clinical outcome measures (see Supplementary Information).

## Discussion

In this study, we show that neurofeedback training with real-time fMRI can reduce the probability that individuals with MDD get stuck in negative attentional states. Our findings indicate that a closed-loop decoding can tap into these states so that they can be externalized for participants by altering the appearance of task stimuli. In doing so, they can recognize when they have slipped into a negative state and gain practice in escaping this state by seeing the fruits of different, often implicit, mental strategies. More broadly, our study demonstrates the feasibility of targeting a theoretically specified cognitive mechanism, decoding its real-time expression from brain activity, and reshaping it through a closed-loop interface.

This neurofeedback approach offers an alternative to classic behavioral therapies targeting attentional biases in MDD (6, 7). Typically, participants are trained to re-orient away from negative stimuli without any read-out of whether the negative stimulus was processed by the brain (1). Our paradigm closes that loop: negative faces become more visible precisely when the classifier indicates that the bias is occurring. Thus, the participant’s training is contingent on transient neural processing of negative content rather than its downstream behavioral output. Moreover, we extend our prior work using this neurofeedback intervention (8) to include an MDD control group, to account for non-specific placebo effects, including expectancy, reward, and task-engagement. Critically, such factors have been associated with improvements in MDD regardless of treatment (9, 10). The comparison between real and sham neurofeedback in randomly assigned individuals with MDD allows us to attribute the reduction in stickiness in the real group to the engagement of a specific process — the coupling between a participant’s own decoded attentional states and the feedback they receive — rather than to repeated behavioral practice in an attentional task alongside rewarding feedback.

In addition to estimating negative attentional biases, we explored how training influenced the neural mechanisms of attention to negative stimuli during the neurofeedback task. Across the whole brain, training decreased activity in the precuneus/PCC and mPFC during periods of high scene visibility. In other words, from the first to the third session, these regions responded less in the real neurofeedback group relative to the sham control group when participants were successfully performing the task (i.e., attending to neutral scenes and ignoring negative faces). One way of interpreting a decrease in precuneus/PCC and mPFC activity is that training reduced the difficulty of the attentional task. Indeed, the mPFC cluster overlaps with the dorsal anterior cingulate cortex (ACC), which has been linked to conflict monitoring (16) and cognitive effort (17, 18). In this view, negative faces may have automatically captured attention at the beginning of training, which required considerable cognitive control to overcome in order to focus on the neutral scenes. By the end of training, the automaticity of this pull toward negative stimuli may have been reduced, thereby lowering conflict and effort.

An alternative explanation is that training increased task engagement. Both the precuneus/PCC and mPFC clusters overlap with the default mode network (DMN), whose suppression is a well-established signature of attentional control (19, 20). A training-related decrease in DMN activity would thus be consistent with improved attentional control in the real group. This interpretation is particularly relevant to MDD. Our mPFC cluster lies between the peaks of two functional subsystems of the DMN: the anterior medial prefrontal core, associated with self-referential processing and the valuation of personally significant information, and the dorsomedial prefrontal subsystem, associated with mentalizing and social-introspective thought (21, 22). Both DMN subsystems have been implicated in the self-referential and ruminative processing characteristic of depression (23), although whether these sub-regions show hyper or hypo activity and connectivity in MDD is task-dependent and inconsistent (24).

Importantly, because the parametric regressor consisted of a single scene vs. face visibility index (scene opacity = 1 - face opacity), the whole-brain effects cannot be attributed unambiguously to task difficulty or even scene processing, rather than to the emotional content of the faces. In other words, the negative relationship between scene visibility and precuneus/PCC and mPFC activity could also be described as a positive relationship with face visibility. We have favored interpretations related to cognitive control out of parsimony, given the well-characterized functions of these regions, but future studies will be needed to better tease out the particular cognitive mechanisms altered by this training. For example, content could be dissociated from difficulty by including blocks with neutral faces superimposed on neutral scenes, as during classifier training (**Figure 1**A), but with variable opacity as in the neurofeedback blocks to track the dynamics of attentional control.

Our findings also relate more generally to theories of the neurocognitive mechanisms underlying MDD. Beck’s cognitive model of depression (25, 26) suggests that schemas instantiated in the brain bias both attention to, and interpretation of, negative information, and that this biased processing leads to a negative feedback loop that underlies core MDD symptoms (26). Moreover, impaired-disengagement models propose that the defining deficit in MDD is not preferential orientation towards negative content but the failure to withdraw attention from it once engaged, a failure attributed to reduced cognitive control over the contents of working memory (4, 27). Our stickiness measure operationalizes this construct, quantifying how often participants remained in the most negative attentional state once they entered it. The reduction in stickiness following real neurofeedback training therefore constitutes evidence that the disengagement deficit is potentially malleable, at least by intervening on the neural correlates of the process if not its behavioral consequences.

Methodologically, our study complements the growing fMRI neurofeedback literature. First, it exemplifies the process-based framework for neurofeedback (28), which suggests that the components of a neurofeedback design should be harmonized around a defined cognitive or affective process rather than the overall classes of symptoms that characterize a mental disorder (like MDD). As such, we target a specific affective process that is deficient in MDD (negative attentional bias), train patients to alter it in the behavioral context it is manifested (via a go/no-go task integrated with closed-loop feedback), and measure its change using an outcome measure tailored to that specific deficit (stickiness measure). Second, our study demonstrates the potential translational value of closed-loop attention training. deBettencourt and colleagues (29) trained sustained attention in healthy adults using real-time decoding of faces vs. scenes to adjust the difficulty of a selective attention task in real-time. Mennen and colleagues (8) applied a similar design to target affective processing, showing that the difficulty MDD patients exhibit in disengaging from negative stimuli can be reduced to levels comparable with healthy controls. The current study uses a double-blind sham-controlled design to show that a reduction in negative attentional biases is specific to genuine neurofeedback, and that this reduction is associated with altered neural processing in evaluative regions linked to MDD. Lastly, rather than training participants to regulate regional, univariate activity, as is typical for real-time fMRI studies in clinical populations, we use a multivariate decoding approach that has become prevalent in basic cognitive neuroscience research to manipulate representational content in the brain (30–33).

Our study has several limitations. First, stickiness increased substantially from the first to the last neurofeedback runs within every session. As a result, we estimated training effects based on a single, initial run per session (run 2) while participants were fresh. Similar within-session decrements have been reported in sustained attention tasks (13) and fMRI neurofeedback studies (12) and most plausibly reflect fatigue accumulating over roughly an hour of a cognitively demanding go/no-go task performed during MRI scanning. Future studies could benefit from dispersing the same number of training runs across more, shorter sessions to reduce fatigue; spreading out training over a larger number of sessions would also provide an increased opportunity for offline, between-session consolidation of neurofeedback benefits (34).

Another limitation is that our main outcome measure was undefined for participants who never entered the most negative attentional bin in run 2 of a particular session. We dealt with this statistically by using a Bayesian model that accounted for missing data in a principled way. Although the model retained 45 of 48 participants, the resulting posterior is wide despite the moderate-to-large effect size. To deal with this issue, future studies could either redefine how stickiness is operationalized to capture negative attentional biases from all participants, or they could include more real neurofeedback runs at the beginning and end of training for both groups, to increase the chance of sampling stickiness in each participant.

Neurofeedback methodology offers several options for control conditions, each with unique advantages and disadvantages (28). Here we chose a yoked-sham control, which matches levels of attention, task engagement, and reward across groups — features associated with placebo effects in MDD (9, 10). However, the lack of contingency between brain activity and sham feedback could potentially worsen performance, for example by providing a false signal that a participant is unable to learn to control, leading them to get discouraged or adopt detrimental strategies. Indeed, we observed a marginally significant increase in stickiness across sessions in the control group (see Supplementary Information). Nonetheless, this increase could also reflect the baseline trajectory of stickiness levels that is not specific to a particular training regime, but rather is the result of the natural history of the deficit (in our case, untreated attentional biases could become worse with time because of fatigue). Thus, the relative difference between real and control groups remains the most incisive way to test for training effects. That said, our main interaction result did not depend only on changes in the control group, as there was also a marginally significant reduction in stickiness across sessions within the real group alone (see Supplementary Information). Future studies could further validate our findings by using a natural history control group or other control group described in (28).

Of note, we did not find group differences in overall symptom severity as measured by MADRS, nor in the behavioral gaze task (see Supplementary Information). These were secondary analyses for which the study was not designed or powered. Our aim was to establish whether the targeted negative attentional bias in the brain can be reduced during training, and whether any such reduction is specific to genuine feedback. Nonetheless, the absence of clinical and behavioral transfer effects leaves open questions about whether the reduced stickiness in our task will generalize to other affective experiences and matter to the lives of individuals with MDD. It is also unclear whether more prolonged training would induce a lasting clinical change and what dose-response curve would govern this change. Future studies could include outcome measures more closely aligned with questionnaires measuring negative attentional biases and cognitive rumination (35, 36), rather than composite MDD severity scales that aggregate across sleep, appetite, mood and psychomotor dimensions.

In sum, our findings indicate that a cognitive bias central to depression can be measured as it unfolds, visually represented to the person expressing it, and reduced through training, with complementary changes in the evaluative circuitry implicated in that bias. Along these lines, as this neurofeed-back approach is refined and validated in future studies and trials, it may provide a causal test of the role of negative attentional biases in depression, and a foundation for precision treatments of this widespread and debilitating disorder.

## Methods

### Preregistration

Preregistration can be found at the ClinicalTrials.gov website (study ID: NCT05169346).

### Participants

Seventy-one participants were screened for the study. Five participants did not qualify based on the inclusion criteria (detailed below). Fifteen participants dropped out either before neurofeedback training was initiated or before completing the three neurofeedback training sessions. Two participants were excluded due to a technical malfunction. One participant was excluded because of erroneous group assignment, leaving 48 participants for analysis (mean age = 29.9, range = [20, 63]; sex = 36 female, 6 male, 6 other; Table S1). Participants were recruited through the University of Pennsylvania Center for Neuromodulation in Depression and Stress and were randomly assigned to real (N=24) or sham (N=24) conditions. All participants attended a total of 6 study visits. The experimental protocol was identical across groups, except for real/sham neurofeedback manipulation, which was conducted in a double-blind fashion. Participants received monetary compensation. The study protocol was approved by the University of Pennsylvania Institutional Review Board and the Princeton University Institutional Review Board through an Institutional Review Board Authorization Agreement.

Participants were recruited with the following inclusion criteria: 18-65 years old; willing to not take psychotropic medications for the duration of the study and not attend psychotherapy sessions; fluent in English; able and willing to provide written consent. To be included in the study, participants had to qualify as having MDD based on the Structured Clinical Interview for DSM-5 Disorders (SCID) questionnaire (37), and to score at least 16 points on the Montgomery-Åsberg Depression Rating Scale (MADRS) clinical interview during Visit 1. After meeting these eligibility requirements, participants were excluded if they: had a medical disorder that may cause depression or require medication that could cause depression symptoms, showed active suicide risk, had MRI contraindications, or were diagnosed with a concurrent DSM-5 psychiatric disorder (e.g., bipolar disorder).

### General procedure

The neurofeedback protocol followed the design of Mennen et al. (8), with modifications described below.

Each participant attended six experimental visits in total. Visits 1, 5 and 6 consisted of a behavior-only assessment at a 1-week and 1-month follow-up, respectively. Where possible, the first 4 visits were scheduled to occur within a 2-week window.

At Visit 1, after obtaining informed consent, participants were administered the SCID (37) to evaluate lifetime and current depressive symptoms and to screen for inclusion criteria. Depression severity over the preceding week was then assessed using the MADRS structured interview (38, 39). The MADRS was repeated at Visits 4 through 6 to track changes in symptom severity over the course of the study.

Neurofeedback training took place over Visits 2 through 4, with participants completing 7 to 9 runs per session (as many as could be accommodated within each 2-hour scanning session). Each run comprised 8 blocks. Blocks in the first run and the first 4 blocks of runs 2 to 9 presented only neutral stimuli at fixed opacity and were used to generate training data for a face vs. scene multivoxel pattern classifier. The remaining 4 blocks (except for run 1) were the neurofeedback blocks, with the target category being neutral scenes and the distractor category being negative faces. During neurofeedback, stimulus opacity was adjusted continuously based on the relative strength of scene versus face classifier evidence as decoded from fMRI activity. Participants were told explicitly that opacity changes reflected their brain activity, not their behavioral performance.

Each block began with a cue specifying both the block type (face or scene) and the target subcategory. For example, a cue reading “outdoor scenes” instructed participants to respond to outdoor scenes (go trials, 90% of trials) and withhold responses to indoor scenes (no-go trials, 10% of trials). Throughout, participants were required to ignore the superimposed, task-irrelevant stimulus category (e.g., faces) while performing the go/no-go judgment. For each participant, one scene subcategory (indoor or outdoor) and one face subcategory (male or female) were designated the “go” target category (the remaining subcategories were “no-go”); this go/nogo subcategory–response mapping was fixed within a participant across sessions and counterbalanced across participants. The order of attend-to-scene versus attend-to-face blocks was likewise counterbalanced across participants, sessions, and runs, whereas the specific image exemplars and their trial order within each block were re-randomized on every run. Each sham participant received a replay of the full stimulus design of their yoked real-group partner.

### Data acquisition

Neuroimaging data were collected using a 3T Siemens Prisma MRI scanner (Siemens AG, Erlangen, Germany) with a 64-channel head coil. Acquisition sequences were matched as closely as possible to those used in Mennen et al. (8) and deBettencourt et al. (29). A high-resolution magnetization-prepared rapid acquisition gradient-echo (MPRAGE) anatomical scan was obtained during the first scanning session and used to construct the whole-brain mask applied both in real time and during offline registration. Registration of the MNI152 standard-space T1-weighted average template to each participant’s functional-space anatomy was performed using FSL (http://fsl.fmrib.ox.ac.uk/), and the resulting whole-brain region of interest served as the mask for that participant. Functional data were acquired using a gradient-echo echo-planar imaging sequence with whole-brain coverage (repetition time = 2 s, echo time = 28 ms, 3-mm isotropic voxels, 64 *×* 64 matrix, 192-mm field of view, 36 slices). Fieldmaps were collected at the end of each session for use in offline processing.

### Double-blinding and randomization

Participants and experimenters were blinded to group allocation throughout the study. Specifically, instructions during training were identical across treatment groups. To ensure experimenter blinding, group assignment was applied automatically via MATLAB code; all analysis steps were identical from the experimenter’s viewpoint throughout data collection. These characteristics rendered the neurofeedback intervention indistinguishable across groups.

### Real-time setup and processing

Real-time neurofeed-back was performed using the RT-cloud software framework (11). During fMRI scanning, each incoming Digital Imaging and Communications in Medicine (DICOM) image was written to a local Linux workstation located in the scanning room. The image was masked, converted into Neuroimaging Informatics Technology Initiative (NIFTI) format and transmitted to a Microsoft Azure cloud server for further preprocessing and classification.

Once the data arrived there, images were spatially smoothed with a Gaussian kernel FWHM of 6 mm. For model training, the classifier was re-trained for each neurofeedback run based on the most recent 8 stable opacity blocks (with neutral faces and 50% opacity for both categories) from before the current run. For example, neurofeedback run 4 used a model trained on the 4 stable blocks from runs 2 and 3. Scikit learn’s logistic regression function (40) was used for model training (solver = ‘saga’, penalty = ‘l2’, max_iter = 300).

Each run’s trained multivoxel pattern classifier (41) was applied during the last 4 neurofeedback blocks to estimate, at each functional time point (TR), the degree to which attention was allocated to the task-relevant scene versus the task-irrelevant face. Each incoming functional image was highpass filtered in real-time (using all time points in the current run up to that point; cutoff = 200 s), and z-scored using the mean and standard deviation from the previous 4 stable blocks of the current run. To provide a single, signed feedback signal — positive when attending to the scene, negative when attending to the face, and zero when the classifier cannot distinguish the two — the face evidence was subtracted from the scene evidence. This difference score was written to a text file and returned to the local Linux machine. This machine controlled the display component of the task using MATLAB (Release 2014a) and Psychtoolbox (42). A script loaded each new text file containing the classifier evidence score once it arrived from the cloud server. The score was converted to an opacity value for the neutral scene using a sigmoidal transfer function. Then, opacity was smoothed using a moving window over the values from the previous two time points to ensure that changes in opacity were not abrupt (29). This smoothed value was set as the scene opacity for the following 3 trials (1.5 TRs) while the next time point was collected and preprocessed; the face opacity was 1 minus the scene opacity.

### Offline MRI preprocessing

All acquired neural data were converted to Brain Imaging Data Structure (BIDS) format (43) and was preprocessed using fMRIPrep 23.2.1 (Esteban et al. (44); Esteban et al. (45); RRID:SCR_016216), which is based on Nipype 1.8.6 (K. Gorgolewski et al. (46); K. J. Gorgolewski et al. (47); RRID:SCR_002502).

The following details below were generated by fMRIPrep and included here as recommended:

T1-weighted images were acquired at the end of the first session. Images were corrected for intensity non-uniformity (INU) with N4BiasFieldCorrection (48), distributed with ANTs 2.5.0 (49), and used as T1w-reference throughout the workflow. The T1w-reference was then skullstripped with a Nipype implementation of the antsBrainExtraction.sh workflow (from ANTs), using OASIS30ANTs as target template. Brain tissue segmentation of cerebrospinal fluid (CSF), white-matter (WM) and gray-matter (GM) was performed on the brain-extracted T1w using fast (RRID:SCR_002823, Zhang, Brady, and Smith 2001). Brain surfaces were reconstructed using recon-all (FreeSurfer 7.3.2, RRID:SCR_001847, Dale, Fischl, and Sereno 1999), and the brain mask estimated previously was refined with a custom variation of the method to reconcile ANTs-derived and FreeSurfer-derived segmentations of the cortical graymatter of Mindboggle (RRID:SCR_002438, Klein et al. 2017). Volume-based spatial normalization to two standard spaces (MNI152Lin, MNI152NLin2009cAsym) was performed through nonlinear registration with antsRegistration (ANTs 2.5.0), using brain-extracted versions of both T1w reference and the T1w template. The following templates were were selected for spatial normalization and accessed with TemplateFlow (50): Linear ICBM Average Brain (ICBM152) Stereotaxic Registration Model [Mazziotta et al. (51); TemplateFlow ID: MNI152Lin], ICBM 152 Nonlinear Asymmetrical template version 2009c [Fonov et al. (52), RRID:SCR_008796; TemplateFlow ID: MNI152NLin2009cAsym].

For each 4D BOLD image per participant (across all sessions), the following preprocessing was performed. First, a reference volume was generated, using a custom methodology of fMRIPrep, for use in head motion correction. Head-motion parameters with respect to the BOLD reference (transformation matrices, and six corresponding rotation and translation parameters) are estimated before any spatiotemporal filtering using mcflirt (53). The estimated fieldmap was then aligned with rigid-registration to the target EPI (echoplanar imaging) reference run. The field coefficients were mapped on to the reference EPI using the transform. The BOLD reference was then co-registered to the T1w reference using bbregister (FreeSurfer) which implements boundary-based registration (54). Co-registration was configured with six degrees of freedom. Several confounding time-series were calculated based on the preprocessed BOLD: framewise displacement (FD), DVARS and three region-wise global signals. FD was computed using two formulations following Power (55) and Jenkinson (53). FD and DVARS are calculated for each functional run, both using their implementations in Nipype (55). The three global signals are extracted within the CSF, the WM, and the whole-brain masks. Additionally, a set of physiological regressors were extracted to allow for component-based noise correction (56). Principal components are estimated after high-pass filtering the preprocessed BOLD time-series (using a discrete cosine filter with 128s cut-off) for the two CompCor variants: temporal (tCompCor) and anatomical (aCompCor). tCompCor components are then calculated from the top 2% variable voxels within the brain mask. For aCompCor, three probabilistic masks (CSF, WM and combined CSF+WM) are generated in anatomical space. The implementation differs from that of Behzadi et al. in that instead of eroding the masks by 2 pixels on BOLD space, a mask of pixels that likely contain a volume fraction of GM is subtracted from the aCompCor masks. This mask is obtained by dilating a GM mask extracted from the FreeSurfer’s aseg segmentation, and it ensures components are not extracted from voxels containing a minimal fraction of GM. Finally, these masks are resampled into BOLD space and binarized by thresholding at 0.99 (as in the original implementation). Components are also calculated separately within the WM and CSF masks. For each CompCor decomposition, the k components with the largest singular values are retained, such that the retained components’ time series are sufficient to explain 50 percent of variance across the nuisance mask (CSF, WM, combined, or temporal). The remaining components are dropped from consideration. The headmotion estimates calculated in the correction step were also placed within the corresponding confounds file. The confound time series derived from head motion estimates and global signals were expanded with the inclusion of temporal derivatives and quadratic terms for each (57). Frames that exceeded a threshold of 0.5 mm FD or 1.5 standardized DVARS were annotated as motion outliers. Additional nuisance timeseries are calculated by means of principal components analysis of the signal found within a thin band (crown) of voxels around the edge of the brain, as proposed by (58). All resamplings can be performed with a single interpolation step by composing all the pertinent transformations (i.e. headmotion transform matrices, susceptibility distortion correction when available, and co-registrations to anatomical and output spaces). Gridded (volumetric) resamplings were performed using nitransforms, configured with cubic B-spline interpolation.

Many internal operations of fMRIPrep use Nilearn 0.10.2 (59), mostly within the functional processing workflow. For more details of the pipeline, see the section corresponding to workflows in fMRIPrep’s documentation.

Finally, pre-processed images were spatially smoothed using a Gaussian kernel at 4-mm FWHM, via nilearn package, version 0.10.2 in Python.

### ROI definition

Two regions of interest (ROIs) were centered on literature coordinates in MNI152NLin2009cAsym space: left amygdala (−18, −8, −16), a 6-mm sphere around a Neurosynth meta-analytic peak for the term “emotional faces” (60); bilateral fusiform face area (FFA), defined based on a probabilistic functional atlas of face-selective regions (61), as the union of two 6-mm spheres, one per hemisphere (left: −40, −54, −20; right: 42, −52, −20). Spheres were resampled with nearest-neighbor interpolation onto the functional BOLD grid to yield the binary masks used by the PPI analysis. The left amygdala sphere served as the seed for the PPI analysis.

### Whole-brain analyses

Analyses were performed in Python using nilearn (59). For each participant and session (1, 3), a first-level general linear model (GLM) was fit to the BOLD data from run 2 after spatial smoothing at 4-mm FWHM. The design included a stable-block regressor modeled as 50-s boxcars, and four separate neurofeedback-block baseline regressors (blocks 5–8), each modeling the mean response of that block separately. A scene vs. face visibility parametric modulator was calculated from the displayed scene vs face opacity proportion as

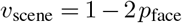

such that (+1) represented a fully visible scene and (−1) represented a fully visible face, and was mean-centered separately within each neurofeedback block. The four block-specific baseline regressors and within-block centering ensured that the parametric modulator captured trial-to-trial variation in scene vs face visibility over and above each block’s mean response. Fixation periods were not modeled explicitly. All task regressors were convolved with the canonical SPM hemodynamic response function (HRF). Low-frequency drift was modeled using discrete-cosine regressors with a 128-s high-pass cutoff, temporal autocorrelation was modeled as an AR(1) process, and each voxel time series was converted to percent signal change. Nuisance regressors included the six rigid-body head-motion parameters, the first six anatomical CompCor components (56) provided by fMRIPrep, and one spike regressor per volume with framewise displacement *≥* 0.2 mm (62).

Participants were excluded from this analysis for excessive head motion if more than 20% of volumes in either analyzed run (run 2 in session 1 and session 3) exceeded the 0.2 mm FD threshold (55, 63). Two participants (with 31.8% and 24.0% of volumes flagged) met this criterion and were excluded, leaving 24 real and 22 sham participants for this analysis.

The contrast of interest was the scene vs face-visibility parametric slope. For each participant, a training-related change map was computed as the voxelwise difference between sessions 3 vs. 1.

Group-level whole-brain inference on the subject-level contrast maps used FSL *randomise* (64) with threshold-free cluster enhancement (TFCE (65)) and 10,000 iterations, restricted to the group functional brain mask. We ran a two-sample test between groups; effects were corrected to *p*(*FWE*) *<* 0.05.

As a manipulation check that the scene vs. face visibility modulation drove the expected category-selective visual processing networks, we pooled the parametric regressor effect across sessions per participant (precision-weighted fixed-effects average) and computed a one-sample t test across all participants, using randomization test with TFCE correction, corrected to *p*(*FWE*) *<* 0.05 (see Supplementary Information).

Task-dependent functional connectivity of the left amygdala was assessed in Python using nilearn (59). We conducted a PPI analysis (66, 67) in the neurofeedback blocks of run 2 in sessions 1 and 3. The physiological regressor was the mean time series extracted from unsmoothed BOLD data within amygdala ROI. The seed time series was adjusted for the nuisance, censoring, and 128-s cosine-drift regressors included in the first-level model, and then demeaned. The psychological variable was the parametric regressor used in the face vs scene visibility analysis, centered separately within each of the four neurofeedback blocks. This variable was modeled at each 1-s trial and convolved with the canonical SPM HRF. The PPI regressor was the product of the cleaned seed amygdala time series and the convolved face-visibility regressor (seed × face visibility). The design additionally included stable-block and fixation regressors, separate baseline regressors for each of the four neurofeedback blocks, a run-wise no-go deviation regressor, and the face-visibility and physiological main effects. The nuisance model (57, 68) comprised the six rigid-body motion parameters, six anatomical CompCor components, and one spike regressor per volume with framewise displacement ≥ 0.2 mm. Temporal autocorrelation was modeled as AR(1). BOLD data were smoothed at 4-mm FWHM and scaled to percent signal change. Participants were excluded from this analysis for excessive head motion if more than 20% of volumes in either analyzed run (run 2 in session 1 and session 3) exceeded the 0.2 mm FD threshold (55, 63). Two participants (with 31.8% and 24.0% of volumes flagged) met this criterion and were excluded, leaving 24 real and 22 sham participants for this analysis.

The first-level contrast of interest was the face-visibility PPI beta map. For each participant, a training-related connectivity-change map was calculated as the session 3 minus session 1 difference in run-2 PPI beta maps.

Whole-brain inference on the subject-level contrast maps was performed with FSL *randomise* (TFCE, 10,000 iterations), restricted to the group functional brain mask, with a two-sample test between groups; maps were corrected at *p*(*FWE*) *<* 0.05. In addition, mean PPI change was extracted from the bilateral FFA ROI, and change was compared between groups using independent t-tests.

## Data Availability

All data produced in the present study are available upon reasonable request to the authors

## ACKNOWLEDGEMENTS

This work was supported by the National Institute of Mental Health of the National Institutes of Health under Award Number [R61MH128492]. The Authors would like to thank Walid Makhoul, Maria Prociuk and Alessia Seroff for their help with data collection.

