## Supplementary Information for "Escaping the Negative Attentional Bias in Depression with Real-Time Neurofeedback"

**References cited in this Supplementary Information are listed in the reference list of the main manuscript.*

### 1. Participant demographics

Demographic characteristics of the 48 analyzed participants (24 real, 24 sham) are presented in Table S1. Sex/gender is self-identified. Race categories are mutually exclusive unless participants report "More than one race"); Hispanic/Latino ethnicity is reported separately.

**Table S1.** Participant demographics by group. Values are $n$ unless stated.

| Characteristic | Real ($n = 24$) | Sham ($n = 24$) | Full cohort ($N = 48$) |
|---|---|---|---|
| *Age (years)* | | | |
| Mean ± SD | $28.8 \pm 9.7$ | $31.0 \pm 8.4$ | $29.9 \pm 9.0$ |
| Range | 21–63 | 20–53 | 20–63 |
| *Gender identity* | | | |
| Woman / female | 19 | 17 | 36 |
| Man / male | 3 | 3 | 6 |
| Non-binary | 1 | 3 | 4 |
| Transgender man (FtM) | 1 | 0 | 1 |
| Transgender woman (MtF) | 0 | 1 | 1 |
| *Race* | | | |
| White | 16 | 13 | 29 |
| Asian | 5 | 3 | 8 |
| Black / African American | 1 | 4 | 5 |
| More than one race | 1 | 4 | 5 |
| Other | 1 | 0 | 1 |
| *Ethnicity* | | | |
| Hispanic / Latino | 1 | 4 | 5 |
| Not Hispanic / Latino | 23 | 20 | 43 |

### 2. Non-parametric test of the group by session interaction in stickiness

The primary interaction (i.e., greater reduction of stickiness in run 2 from session 1 to session 3 in real group than sham group) was estimated with a Bayesian hierarchical model retaining all participants with at least one timepoint (23 real, 22 sham). To confirm that the result did not depend on the model's distributional assumptions or its handling of missing data, we re-tested it with non-parametric analyses of complete-case data; that is, in participants (16 real, 12 sham) with a stickiness score in run 2 from both session 1 ("pre") and session 3 ("post").

We first ran a Wilcoxon rank-sum test on the pre→post change. For each participant, we computed the post minus pre difference (negative = reduction) and compared it between groups. Real neurofeedback training significantly reduced the probability of getting stuck in the most negative state compared to the yoked sham control (real mean $= -0.107$, sham mean $= 0.207$; Wilcoxon rank-sum: $W(U) = 48$, $z = -2.228$, two-sided $p = 0.027$; rank-biserial effect size = 0.5).

We complemented the rank-sum test, which does not assume normality but discards magnitude, with a non-parametric bootstrap analysis to estimate the group by session interaction. This analysis keeps the original scale, matches the mean-level statistic of the primary Bayesian model, and yields a confidence interval. Each complete-case participant had one post minus pre change score, so resampling was at the participant level. On each of 50,000 iterations, we sampled with replacement and within each group separately the total number of change scores in that group (16 real, 12 sham); we then calculated the interaction term as the difference of the within-group mean change scores (real minus sham), together with an accompanying effect size equal to this double difference divided by the pooled standard deviation of the sampled change scores in each group (a bootstrap Cohen's $d$). The resulting 50,000 interaction values formed the sampling distribution from which we obtained the 95% percentile confidence intervals, the directional $p$ (proportion of interations with the interaction value $\geq 0$), and the median effect size. This bootstrap approach reproduced the key interaction result (mean interaction value $= -0.31$, 95% CI $[-0.58, -0.04]$, $p = 0.013$, median bootstrap Cohen's $d = -0.94$, CI $[-2.02, -0.10]$; Figure S1).

The rank-based and mean-based non-parametric tests yield nearly identical results and both agree with the primary Bayesian interaction: a greater reduction of stickiness in the most negative attentional state across training in the real group more than the sham group.

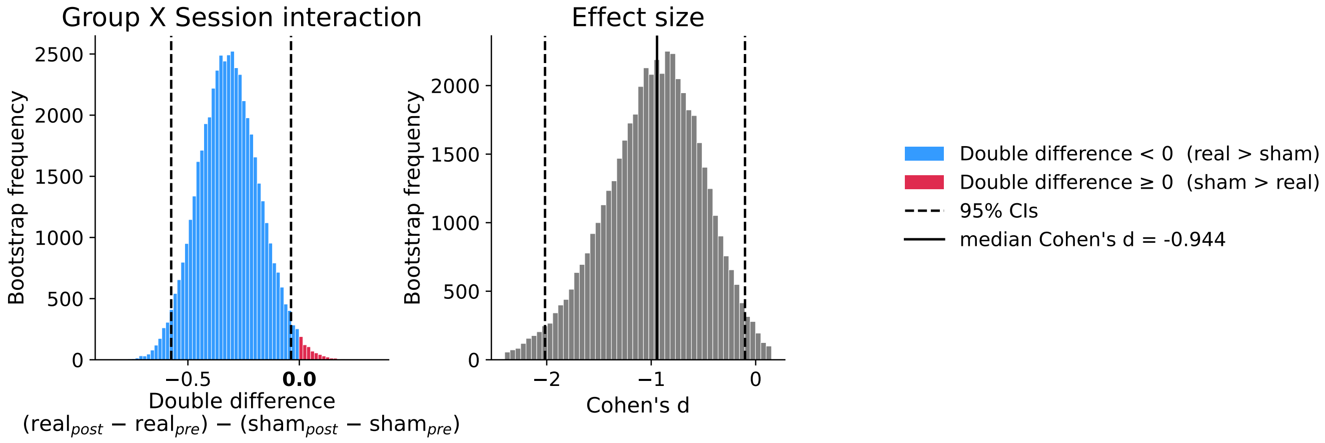


**Fig. S1.** Non-parametric bootstrap of the group by session interaction. Left: bootstrap distribution of the interaction double difference (blue mass below zero indicates larger reduction in the real group; the $\geq 0$ mass is the directional $p$-value). Right: bootstrap distribution of the interaction effect size (Cohen's $d$). Dashed lines, 95% CIs.

#### 3. Within-group pre to post changes in stickiness

The key outcome measure in the main text and above is the group by session interaction. This interaction, via the logic of a randomized controlled design, allows us to conclude that real neurofeedback was effective at reducing stickiness above and beyond any generic effect of time or practice. As a follow-up analysis, we asked whether stickiness changed from pre to post separately within each group. We report both a non-parametric bootstrap analysis of complete-case data, which uses only participants with both pre and post data and makes no assumption about the missing data, as well as a hierarchical Bayesian model, which can recover partially observed participants by imputing a missing timepoint.

The bootstrap analysis was performed similarly to the interaction test above, except that a sampling distribution of the post minus pre change score was generated separately for each group. The real group ($n = 16$ complete participants) showed a trend toward decreased stickiness from pre to post (mean pre $= 0.21$, mean post $= 0.10$; mean change $= -0.11$, 95% CI $[-0.24, 0.03]$, mean Cohen's $d = -0.38$, $p = 0.059$). The sham group ($n = 12$) increased over the same interval (mean pre $= 0.17$, mean post $= 0.38$; mean change $= 0.21$, 95% CI $[-0.03, 0.43]$, mean Cohen's $d = 0.49$, $p = 0.044$).

The Bayesian model was fit separately for each group with a per-subject random intercept and weakly informative priors on the pre-to-post change (non-centered parameterization; 4 chains $\times$ 4,000 draws; all $\hat{R} \leq 1.01$, no divergences). Partially observed subjects were retained by imputing the missing timepoint under the likelihood. The pattern of results was the same as the bootstrap analysis but with smaller effects, with numerically decreased stickiness in the real group ($n = 23$, mean change $= -0.07$, 95% HDI $[-0.20, 0.06]$, posterior probability of decrease $P(\text{change} < 0) = 0.87$) and numerically increased stickiness in the sham group ($n = 22$, mean change $= 0.16$, 95% HDI $[-0.04, 0.35]$, posterior probability of increase $P(\text{change} > 0) = 0.95$; mean standardized effects $-0.38$ and $0.57$, respectively). The smaller effects relative to the bootstrap analysis could stem from the partially observed subjects, recovered here by imputation, pulling each group's estimate toward zero, weakening the directional evidence.

#### 4. Whole-brain manipulation check for face and scene processing

As a manipulation check, we tested whether the varying visibility of scene vs. face images during neurofeedback drove corresponding category-selective neural responses. We expected high face visibility to increase activity in face-selective visual regions, such as the fusiform face area (FFA), and high scene visibility to increase activity in scene-selective visual regions, such as the parahippocampal place area (PPA). For each participant, we averaged the first-level contrast maps of the face vs. scene parametric regressor effect across sessions 1 and 3 using a precision-weighted fixed-effects average, yielding one map per participant. Whole-brain group effects across participants were estimated using a randomization test (FSL *randomise*; 10,000 permutations), and corrected for multiple comparisons using threshold-free cluster enhancement (TFCE; p(FWE)<0.05).

Analysis confirmed that scene vs. face visibility drove expected neural responses (Figure S2). Negative effects (purple), corresponding to high face visibility, were concentrated in bilateral ventral occipitotemporal cortex, including the FFA. Positive (green) effects, corresponding to high scene visibility, showed widespread engagement of medial occipitotemporal cortex extending to the PPA and retrosplenial cortex, and to a broader network that co-activates with these regions (69), including in lateral parietal cortex and mPFC.

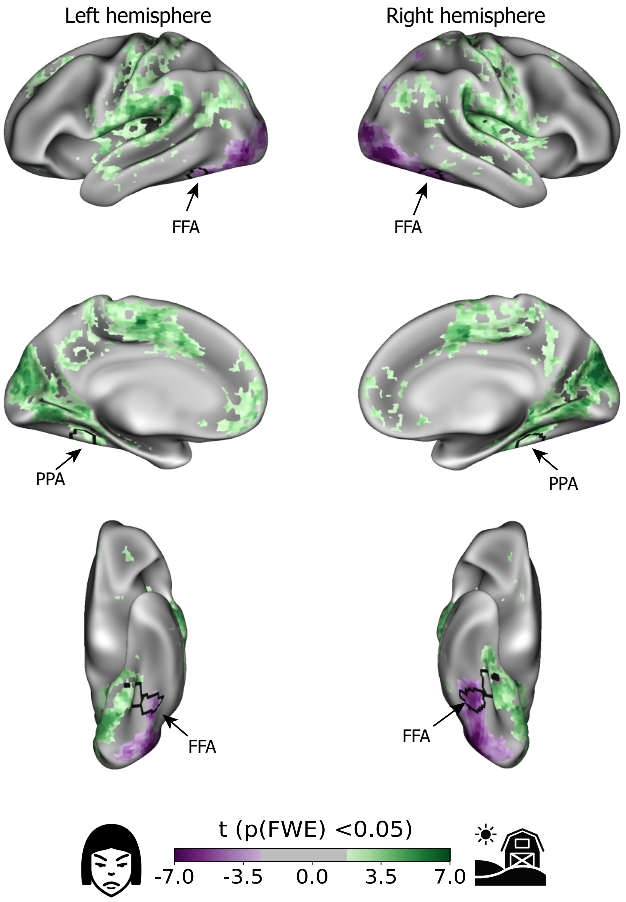


**Fig. S2.** Whole-brain parametric map of the scene vs. face visibility averaged across sessions. Green and purple colors indicate significant positive and negative clusters, respectively (TFCE, $p_{\mathrm{FWE}} < .05$). Black contours marked with FFA and PPA are 6-mm spheres centered on peak coordinates from prior studies (61, 70). Maps are displayed on inflated surfaces in lateral (top), medial (middle), and ventral (bottom) views.

### 5. Behavioral transfer to free-viewing gaze task

To test whether neurofeedback training transferred to spontaneous attention allocation during free viewing, participants completed an eye-tracking task at the baseline visit and after each of the three neurofeedback sessions. The task was adapted from Mennen et al. (8) and, originally, from Kellough et al. (71). On each trial, four images — one each of dysphoric, threatening, happy, and neutral — were displayed simultaneously in different quadrants for 30 s while participants viewed the screen freely (instructed to look "as if watching television"). Eye position was recorded at 120 Hz. Each session comprised 20 trials in random order: 12 valence trials (the four-valence array) and 8 neutral-filler trials (four neutral images). Stimuli were drawn from the International Affective Picture System (72), the Nencki Affective Picture System (73), and the original paradigm. They were arranged into four session-specific versions matched on image entropy, contrast, and luminance (version order randomized across participants).

Raw gaze samples were converted into fixations (runs of $\geq 100$ ms or $\geq 12$ consecutive samples at 120 Hz that remained within $1^\circ$ of visual angle) and each fixation was assigned to a quadrant. The number of usable fixations varied across trials and participants (e.g., because of blinks, glasses, eye make-up), so every metric was normalized by the total number of recorded fixations. For each emotional category, we computed four metrics per session: (i) *initial orientation*: the probability that a trial's first fixation landed on that category; (ii) *total viewing time*: the absolute time (ms) spent fixating on that category; (iii) *duration ratio*: viewing duration of a certain category divided by the trial's total fixation; and (iv) *maintenance ratio*: the fraction of fixation sustained on a category once it was first fixated, indexing difficulty disengaging. Trial-level values were averaged to one value per category, session, and participant.

For each of the four metrics in the four emotional categories (16 tests), the per-participant values were entered into a mixed-model ANOVA with group (real vs. sham) as the between-subjects factor and session (baseline, 1, 2, 3) as the within-subjects factor, restricted to participants for with complete data ($N = 45 - 47$ depending on the metric and emotion). Behavioral transfer from neurofeedback training was estimated by the group by session interaction, which did not reach significance for any test (Table S2).

**Table S2.** Group by session interaction by metric and emotion in the gaze task.

| Metric | Emotion | $F$(df1, df2) | $p$ | $\eta_p^2$ | $N$ |
|---|---|---|---|---|---|
| Maintenance Ratio | Dysphoric | 0.62 (3, 135) | 0.601 | 0.014 | 47 |
| Maintenance Ratio | Threat | 0.19 (3, 135) | 0.905 | 0.004 | 47 |
| Maintenance Ratio | Neutral | 0.73 (3, 135) | 0.533 | 0.016 | 47 |
| Maintenance Ratio | Positive | 0.29 (3, 135) | 0.834 | 0.006 | 47 |
| Duration Ratio | Dysphoric | 1.02 (3, 132) | 0.386 | 0.023 | 46 |
| Duration Ratio | Threat | 0.72 (3, 132) | 0.542 | 0.016 | 46 |
| Duration Ratio | Neutral | 1.05 (3, 132) | 0.374 | 0.023 | 46 |
| Duration Ratio | Positive | 0.70 (3, 132) | 0.551 | 0.016 | 46 |
| Total time (ms) | Dysphoric | 1.85 (3, 129) | 0.141 | 0.041 | 45 |
| Total time (ms) | Threat | 0.19 (3, 129) | 0.904 | 0.004 | 45 |
| Total time (ms) | Neutral | 0.48 (3, 129) | 0.696 | 0.011 | 45 |
| Total time (ms) | Positive | 1.24 (3, 129) | 0.299 | 0.028 | 45 |
| Initial Orientation | Dysphoric | 0.25 (3, 135) | 0.860 | 0.006 | 47 |
| Initial Orientation | Threat | 1.46 (3, 135) | 0.229 | 0.031 | 47 |
| Initial Orientation | Neutral | 0.48 (3, 135) | 0.698 | 0.011 | 47 |
| Initial Orientation | Positive | 0.83 (3, 135) | 0.482 | 0.018 | 47 |

### 6. Clinical transfer to MADRS

Participants completed the MADRS (38) before training (baseline) and after each neurofeedback session (sessions 1–3), plus follow-ups after 1 week and 1 month. We averaged the two follow-up MADRS scores for each participant into an overall "mean-FU" score (participants with only one follow-up contributed that single value). MADRS scores were entered into a mixed-model ANOVA with group (real vs. sham) as the between-subjects factor and timepoint (baseline, mean-FU) as the within-subjects factor, restricted to participants with a baseline score and at least one follow-up ($N = 46$: 24 real, 22 sham). There was a significant main effect of timepoint ($F(1,44) = 38.2$, $p < 0.0001$, $\eta_p^2 = 0.47$) but no main effect of group ($F(1,44) < 0.01$, $p = 0.99$, $\eta_p^2 < 0.001$) and no group by timepoint interaction ($F(1,44) = 0.23$, $p = 0.635$, $\eta_p^2 = 0.005$). Therefore, we found no evidence of differential symptom change between the real and sham groups (Table S3).

**Table S3.** MADRS score (mean $\pm$ SD) by group and timepoint.

| Group | Baseline | 1-week follow-up | 1-month follow-up | Mean-FU |
|---|---|---|---|---|
| Real ($n = 24$) | $27.9 \pm 5.8$ | $21.7 \pm 9.0$ | $19.4 \pm 9.1$ | $20.7 \pm 8.2$ |
| Sham ($n = 22$) | $28.4 \pm 3.7$ | $21.2 \pm 9.2$ | $19.7 \pm 9.1$ | $20.1 \pm 8.7$ |

We assessed the correlation between change in stickiness (session 3 − 1 for run 2) and change in MADRS (mean-FU − baseline), collapsed across groups ($N = 27$) and within each group (16 real, 11 sham; one sham participant with complete-case neurofeedback did not attend the post-training follow up sessions). No significant association was observed (real: $r = 0.05$, $p = 0.857$; sham: $r = 0.20$, $p = 0.547$).
